# Needs Assessment for the Development of a Sustainability Curriculum for Surgical Residents

**DOI:** 10.1101/2024.05.15.24307424

**Authors:** Madeline Matthys, Jane Wang, Tejas S. Sathe, Kaiyi Wang, Seema Gandhi, Hanmin Lee, Adnan Alseidi, the Sustainability Curriculum Working Group

**Affiliations:** Department of Surgery, University of California, San Francisco, San Francisco, CA; Department of Anesthesia, University of California, San Francisco, San Francisco, CA

**Author notes:** **Corresponding Author** Madeline Matthys, BS, Department of Surgery, University of California, San Francisco, 505 Parnassus Ave, San Francisco, CA 94143. **Sustainability Curriculum Working Group Group Authorship:** Alexandra Bourdillon, MD - Department of Otolaryngology – Head and Neck Surgery, University of California, San Francisco, San Francisco, CA Riley Brian, MD, MAEd - Department of Surgery, University of California, San Francisco, San Francisco, CA Lucia Calthorpe, MD, MPhil - Department of Surgery, University of California, San Francisco, San Francisco, CA Simon N. Chu, MD, MS - Department of Surgery, University of California, San Francisco, San Francisco, CA Edwin Eshaghzadeh, MD, DDS - Department of Oral and Maxillofacial Surgery, University of California, San Francisco, San Francisco, CA Kara Faktor, MD, MSc - Department of Surgery, University of California, San Francisco, San Francisco, CA Jasmine Huang, MD - Department of Surgery, Lehigh Valley Health Network, Allentown, PA Amandine Godier-Furnemont, MD - Department of Surgery, University of California, San Francisco, San Francisco, CA Sarah Lund, MD - Department of Surgery, Mayo Clinic, Rochester, MN Wendelyn Oslock, MD, MBA - Department of Surgery, University of Alabama at Birmingham, Birmingham, AL Thomas Sorrentino, MD - Department of Surgery, University of California, San Francisco, San Francisco, CA Nichole Starr, MD, MPH - Department of Surgery, University of California, San Francisco, San Francisco, CA Ellen Tsay, MD - Department of Orthopaedic Surgery, University of California, San Francisco, San Francisco, CA Ava Yap, MD, MHS - Department of Surgery, University of California, San Francisco, San Francisco, CA Catherine N. Zivanov, MD - Department of Surgery, Washington University in St. Louis, St. Louis, MO. **Generative AI** No generative AI was used in the writing of this manuscript.

**Keywords:** Surgical Sustainability, Surgical Education, Sustainability Curriculum, Needs Assessment, Graduate Medical Education

## Abstract

**Introduction:** The healthcare sector accounts for 8.5% of United States (U.S.) greenhouse gas emissions, of which one-third comes from operating rooms (ORs). As a result, there is great interest in decarbonizing the OR and surgical care. However, surgical residents are not routinely educated on the negative environmental impact of surgery or how to reduce it. In this paper, we present a formal needs assessment for a sustainability curriculum geared towards surgical residents.

**Methods:** Using Kern’s Six-Step Framework for curriculum development, we conducted focus groups with surgical residents to perform a targeted needs assessment on three main topics: 1) the current state of surgical sustainability curricula; 2) resident knowledge regarding the environmental impact of surgery and barriers to sustainable practice; and 3) preferred educational methods and topics within sustainability education. We recorded all focus groups and performed thematic analysis using anonymized transcripts.

**Results:** Fourteen residents participated in three focus groups, from which a qualitative analysis revealed four themes. First, surgery residents receive limited formal teaching on the negative environmental impact of surgical care or how to reduce this impact. Second, surgery residents have variable levels of prior education about and interest in sustainability in surgery. Third, several barriers prevent the implementation of sustainable changes in surgical practice, including a lack of institutional initiative, cultural inertia, concerns about workflow efficiency, and limited formal education. Finally, residents prefer to learn about practical ways to reduce waste, specifically through interactive approaches such as quality improvement initiatives.

**Conclusions:** Given the increasing importance of sustainability in surgery, there is an urgent need for formal resident education on this topic. This needs assessment provides a valuable foundation for future sustainability curriculum development.

## Introduction

Education regarding operating room sustainability is infrequently offered and prioritized, with over 70% of perioperative faculty and staff reporting a lack of formal curricula.^1^ However, the urgency of the climate crisis requires that surgical departments and training programs prioritize this increasingly important topic. The United States (U.S.), among many other nations, has publicly committed to reducing greenhouse gas emissions to 50% below 2005 levels by 2030 and achieving net-zero emissions by 2050.^2^ These targets represent the levels of decarbonization required to cap the rise in global temperatures to less than two degrees Celsius above pre-industrial levels, an amount that would be catastrophic to human health.^3^ As the healthcare sector accounts for 8.5% of U.S. greenhouse gas emissions,^4^ it is clear that reducing healthcare emissions could dramatically decrease the nation’s carbon footprint. Within the healthcare sector, operating rooms are three to six times more energy-intensive per unit area than the hospital as a whole.^5^ As a result, engaging current and future surgeons in healthcare decarbonization efforts is essential.

Although awareness about sustainability is increasing in academic surgery, significant variability exists in sustainability education, interest in decarbonization, and buy-in at the individual and institutional levels.^1^ This may stem from a lack of awareness regarding the amount of waste operating rooms generate, how this waste contributes to climate change, or what individuals can do to reduce it. In fact, in a recent survey of academic surgeons, a lack of education was cited as a barrier to improving sustainability.^6^ To date, no study has evaluated the need for or the ideal structure of a sustainability curriculum for surgical residents.

The goal of our study was threefold: first, to understand the degree to which surgical residents are exposed to formal curricula regarding sustainability in surgery; second, to identify resident beliefs and attitudes regarding sustainable surgical practices and education; and third, to understand if and how surgical residents want to learn about sustainability.

## Methods

### Focus Group Design and Completion

Following Kern’s Six-Step Framework for curriculum development, two general surgery residents (JW, TS) and one fourth-year medical student (MM) conducted a targeted needs assessment through focus groups comprised of surgical residents. One surgery faculty mentor (AA) and one anesthesia faculty mentor (SG) advised the team, as they both have significant medical education, surgical education, and curriculum development backgrounds. We conducted this study in accordance with the COREQ guidelines.^7^

We constructed a focus group guide containing the main topics and guiding questions, iteratively refined this guide based on feedback from faculty mentors (AA and SG), and piloted the focus group questions with one member of the Sustainability Curriculum Working group (AG-F). This resident provided feedback on format and did not participate in the formal focus groups. For the focus groups, we used a semi-structured group interview format followed by group discussion. Final topics and guiding questions used to lead the focus groups are listed in Table 1.

**Table 1.** Outline of overarching topics and associated guiding questions for focus groups.

| Objectives | Focus Group Questions |
| --- | --- |
| **Objective 1**<br>Assess residents’ current knowledge and awareness of sustainability and waste reduction in the operating | What education (if any) do surgical residents currently receive about sustainability, waste reduction, and carbon footprint of surgical spaces and perioperative spaces?<br>What would motivate surgical residents to educate themselves on surgical sustainability? |
| room, as well as the extent of sustainability education in surgical training. | How does climate change impact patient health and/or outcomes?<br>Can you share any experiences or observations related to waste and/or reducing waste in the operating room?<br>What challenges or barriers do you think may exist when implementing sustainable practices in the operating room? |
| **Objective 2**<br>Identify resident attitudes, beliefs, and concerns regarding sustainable surgical practices and education. | How would a surgical sustainability curriculum fit into the existing constraints with working 80 a week, didactics, studying, etc.? |
| **Objective 3**<br>Gather insights on the preferred learning methods and areas of interest for a sustainability-focused curriculum. | Of the following topics, how would you rank them in terms of interest and usefulness? What other topics would you add?<br>1. OR carbon footprint impacts on global climate change (knowledge-based)<br>2. Patient impact from climate change (knowledge-based)<br>3. Methods to reduce OR waste (implementation)<br>What learning methods (e.g., lectures, workshops, simulations, online modules) do you believe would be most effective for teaching sustainable surgical practices? |

#### Participants

We used voluntary sampling via email and social media to recruit U.S. residents from multiple post-graduate years (PGY) and surgical specialties, including general surgery, orthopedic surgery, otolaryngology (ENT), and oral and maxillofacial surgery (OMFS). As compensation, we provided a meal for all in-person participants. All participants were also offered group authorship on the resulting manuscript and placed in a raffle for a $100 gift card. At the start of each focus group, participants provided verbal consent for audio recording. This study was approved by the University of California, San Francisco’s (UCSF) Institutional Review Board (IRB 23-39152).

### Data Analysis and Validation

We recorded audio for each focus group and used Otter.ai (Otter.ai, Inc., Mountain View, CA) to transcribe each recording. MM manually anonymized the transcriptions to remove any identifying information. Authors used Dedoose Version 9.0.107 (Dedoose, Manhattan Beach, CA) for coding and thematic analysis of the anonymized transcripts. Following the steps laid out by Kiger et al.,^8^ MM, JW, and TS manually reviewed each transcript to individually formulate codes and then met as a group to finalize the codebook. The three authors then individually posited themes from these codes to create thematic maps, and again met as a group to consolidate the final thematic results.

### Reflexivity

Our research team considered reflexivity throughout data collection (focus groups), inductive data analysis, and thematic synthesis.^9^ The members involved in the design and analysis included a medical student (MM), surgical residents (JW, TS), and faculty members from different departments including hepatopancreaticobiliary surgery (AA) and anesthesia (SG). These members had backgrounds in both surgical education and educational theory, which allowed us to incorporate diverse perspectives into the formulation and analysis of focus groups.

## Results

### Demographics

We conducted three one-hour focus group sessions and interviewed fourteen residents across the three sessions: seven in the first in-person focus group, three in the second in-person focus group, and four in the virtual focus group. Surgical residents from all levels of training participated in the focus groups, though the majority (35%) were research residents. Eleven participants (79%) were general surgery residents, including all four virtual focus group participants. The remaining three participants included one orthopedic surgery resident, one ENT resident, and one OMFS resident (Table 2). The majority (71%) of residents included in the focus group were from UCSF, a prominent academic institution located in California.

**Table 2:** Focus Group Demographics.

|  | % (n) |
| --- | --- |
| **Gender** |  |
| Female | 71% (10) |
| Male | 29% (4) |
| Non-binary | 0% (0) |
| **Level of Training** |  |
| PGY1 | 14% (2) |
| PGY2 | 7% (1) |
| PGY3 | 14% (2) |
| PGY4 | 21% (3) |
| PGY5 | 7% (1) |
| Research Year Resident | 36% (5) |
| **Specialty** |  |
| General Surgery | 19% (11) |
| Oral and Maxillofacial Surgery | 7% (1) |
| Orthopedic Surgery | 7% (1) |
| Otolaryngology | 7% (1) |

### Thematic Analysis

We identified four themes within the focus group data. Each theme along with representative quotations is provided below. Table 3 contains a complete list of themes, sub-themes, and additional representative quotes.

**Table 3:**
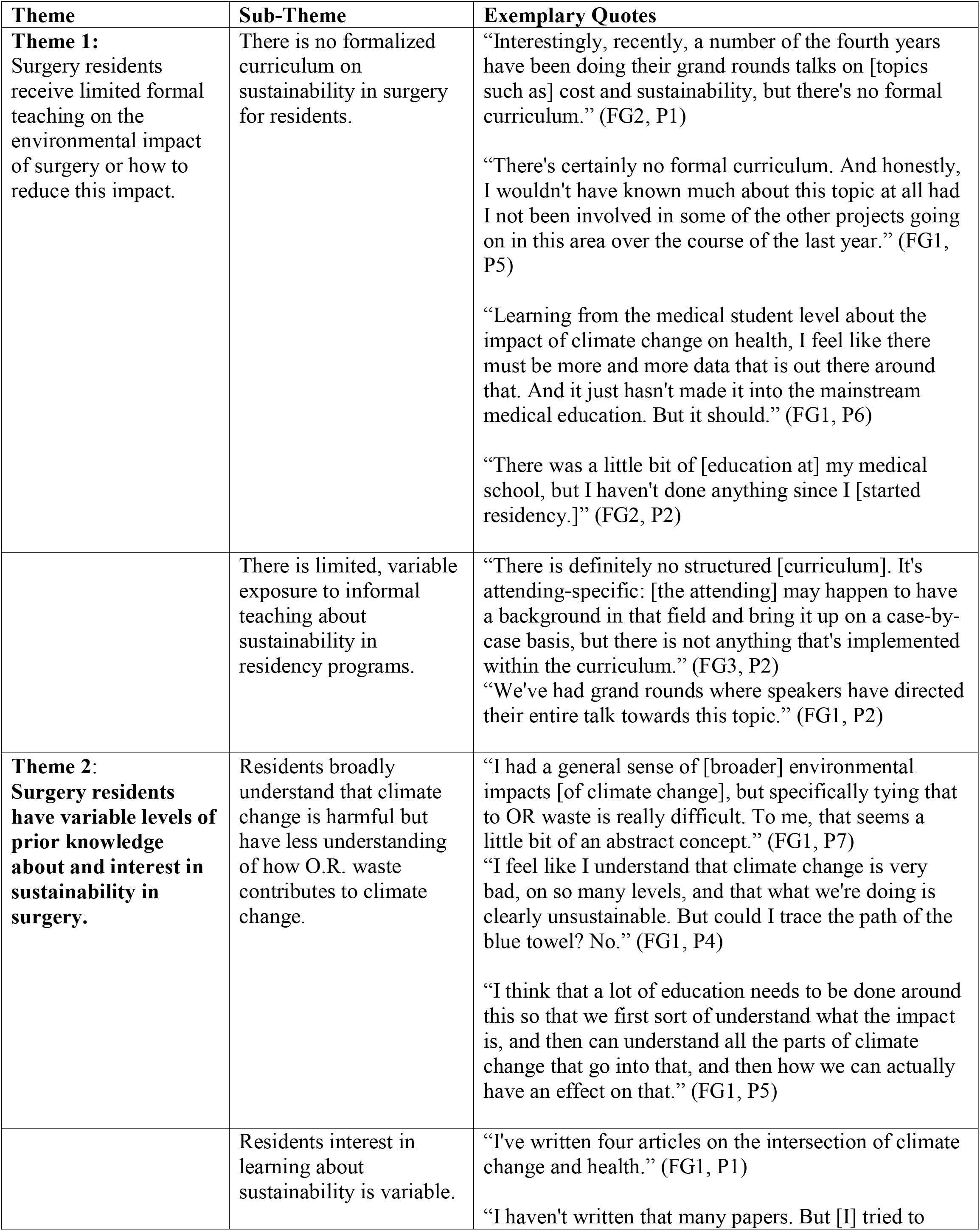
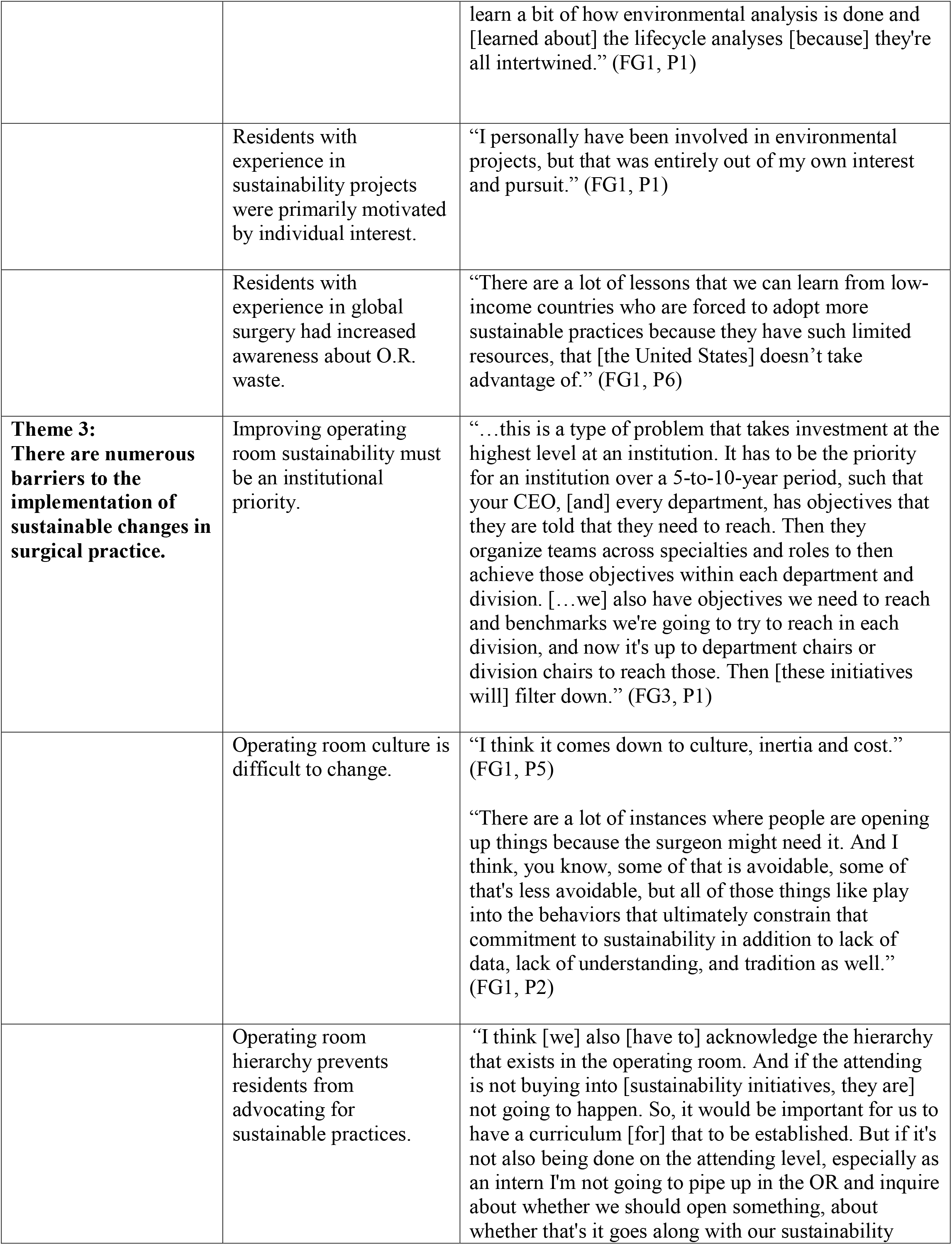
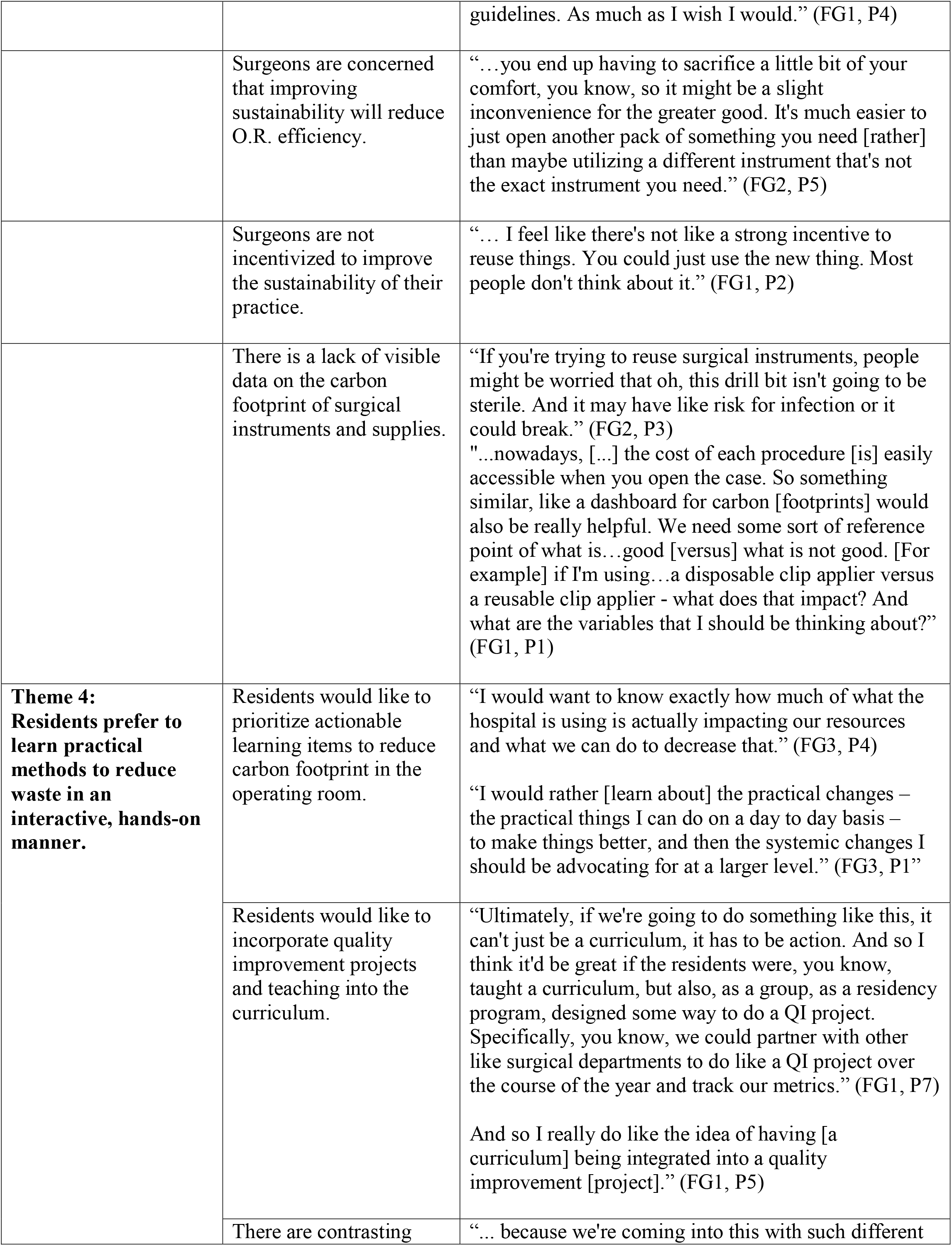
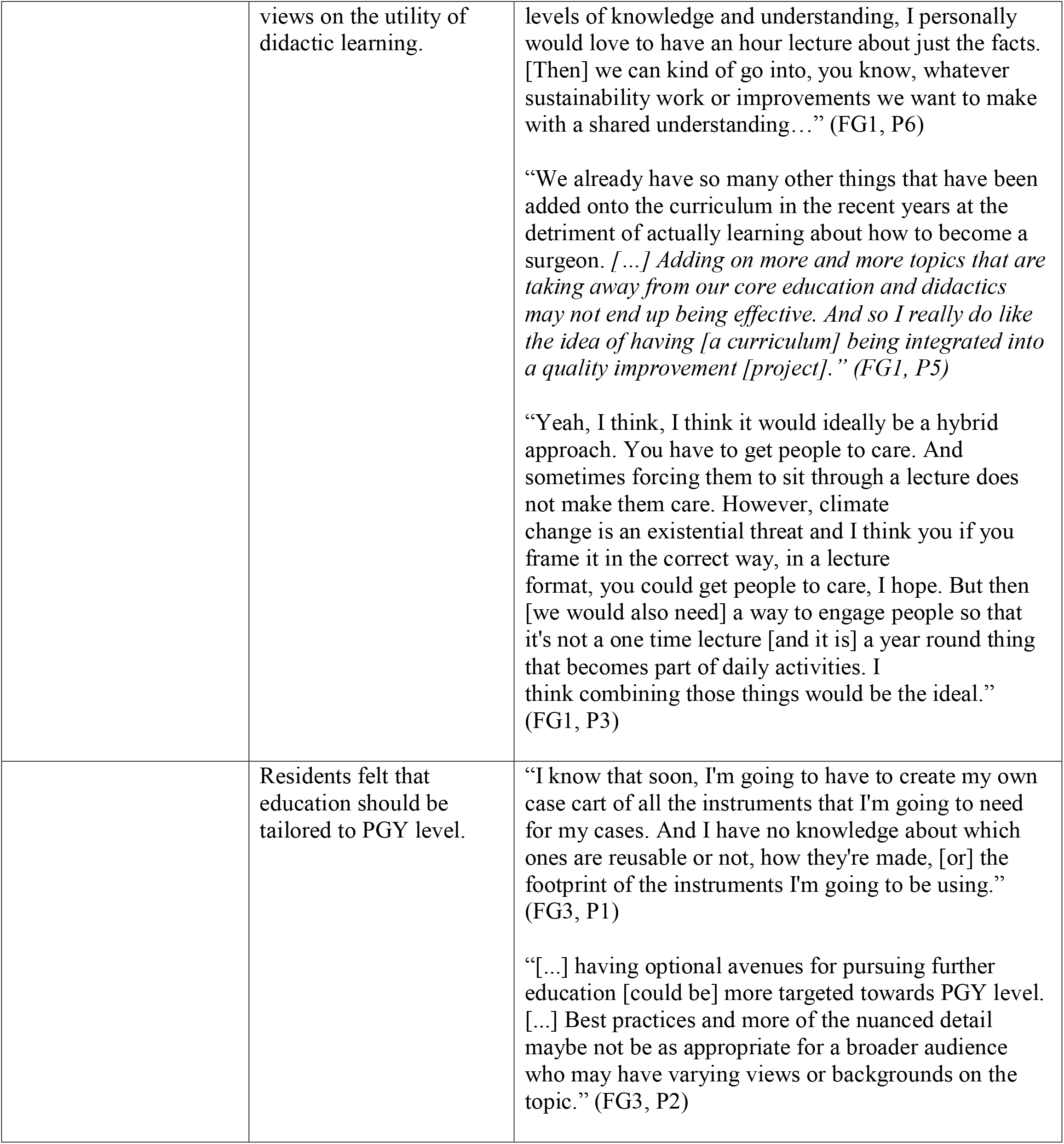
Themes and sub-themes identified through analysis that were associated with each framing topic, as well as exemplary quotes associated with each.

#### Theme 1: Surgery residents receive limited formal teaching on the negative environmental impact of surgery or how to reduce this impact

Residents reported that they received limited to no formal curricula regarding how healthcare contributes to climate change, the impact of climate change on patients, or how to make the operating room more environmentally sustainable.

> *“Interestingly, recently, a number of the fourth years have been doing their grand rounds talks on [topics such as] cost and sustainability, but there’s no formal curriculum*.*” (FG2, P1)*
>
> *“There’s certainly no formal curriculum. And honestly, I wouldn’t have known much about this topic at all had I not been involved in some of the other projects going on in this area over the course of the last year*.*” (FG1, P5)*

At present, there are limited avenues through which residents hear about this topic in a hospital setting, as explained below.

> *“There is definitely no structured [curriculum]. It’s attending-specific: [the attending] may happen to have a background in that field and bring it up on a case-by-case basis, but there is not anything that’s implemented within the curriculum*.*” (FG3, P2)*

#### Theme 2: Surgery residents have variable levels of prior education about and interest in sustainability in surgery

Despite the lack of formal training, most residents understood that climate change is a global threat. These residents referenced natural disasters and extreme weather events when asked how climate change impacts patients. However, they were infrequently able to point out less direct impacts of climate change, such as food insecurity from increased drought conditions. Moreover, residents were cognizant that operating rooms generate significant amounts of waste but lacked a rigorous framework to quantify this waste and its environmental impact. Notably, most residents did not have an understanding of how operating room waste or supply chains are managed at an institutional level.

> *“I had a general sense of [broader] environmental impacts [of climate change], but specifically tying that to OR waste is really difficult. To me, that seems a little bit of an abstract concept*.*” (FG1, P7)*

Resident interest in the topic of sustainability in surgery was variable. Among our respondents, we had some that only heard about sustainability passively, some that pursued scholarly work in sustainability, and others who led projects to improve sustainability such as one resident who contributed to a pilot study implementing reusable gowns.^10^

> *“I’ve written four articles on the intersection of climate change and health*.*” (FG1, P1)*
>
> *“I haven’t written that many papers. But [I] tried to learn a bit of how environmental analysis is done and [learned about] the lifecycle analyses [because] they’re all intertwined*.*” (FG1, P1)*

Specifically, residents with experience in sustainability-related work were primarily motivated by interest in the environment.

> *“I personally have been involved in environmental projects, but that was entirely out of my own interest and pursuit*.*” (FG1, P1)*

Of note, several residents with experience in resource-limited settings, such as in global surgery, expressed greater awareness of operating room waste and the importance of sustainability. Notably, these residents viewed sustainability as a necessity in the setting of resource scarcity rather than just an intrinsic good, as described below:

> *“There are a lot of lessons that we can learn from low-income countries who are forced to adopt more sustainable practices because they have such limited resources, that [the United States] doesn’t take advantage of*.*” (FG1, P6)*

#### Theme 3: There are numerous barriers to the implementation of sustainable changes in surgical practice

When asked about potential barriers to the implementation of sustainable practices, several residents pointed to a lack of institutional buy-in. Overall, there was skepticism that sustainable practices could be implemented at scale in a grassroots manner. As one resident said,

> *“…this is a type of problem that takes investment at the highest level at an institution. It has to be the priority for an institution over a 5-to-10-year period, such that your CEO, [and] every department, has objectives that they are told that they need to reach. Then they organize teams across specialties and roles to then achieve those objectives within each department and division. […we] also have objectives we need to reach and benchmarks we’re going to try to reach in each division, and now it’s up to department chairs or division chairs to reach those. Then [these initiatives will] filter down*.*” (FG3, P1)*

Residents also felt that cultural stagnancy with certain practices, the inherent hierarchy of the operating room, and an emphasis on efficiency made it difficult to voice concerns or challenge the status quo.

> *“I think [we] also [have to] acknowledge the hierarchy that exists in the operating room. And if the attending is not buying into [sustainability initiatives, they are] not going to happen*.*” (FG1, P4)*

In addition, residents noted the lack of incentives or frequent reminders to make individual sustainable choices. One resident compared this to the fact that data regarding the cost of individual instruments are readily available on electronic medical records prior to and following cases. In contrast, similar data regarding the carbon footprint of each item are less available.

> *“… I feel like there’s not like a strong incentive to reuse things. You could just use the new thing. Most people don’t think about it*.*” (FG1, P2)*
>
> *“*…*nowadays, [*…*] the cost of each procedure [is] easily accessible when you open the case. So something similar, like a dashboard for carbon [footprints] would also be really helpful. We need some sort of reference point of what is…good [versus] what is not good. [For example] if I’m using…a disposable clip applier versus a reusable clip applier - what does that impact? And what are the variables that I should be thinking about?” (FG1, P1)*

#### Theme 4: Residents prefer to learn practical methods to reduce waste in an interactive manner

Residents primarily wanted to learn about actionable steps they could take to reduce their carbon footprint in the hospital

> *“I would want to know exactly how much of what the hospital is using is actually impacting our resources and what we can do to decrease that*.*” (FG3, P4)*
>
> When asked how the curriculum should be structured, most residents preferred an interactive curriculum focused on practical quality improvement initiatives, as described below.
>
> *“Ultimately, if we’re going to do something like this, it can’t just be a curriculum, it has to be action. And so I think it’d be great if the residents were, you know, taught a curriculum, but also, as a group, as a residency program, designed some way to do a QI project. Specifically, you know, we could partner with other like surgical departments to do like a QI project over the course of the year and track our metrics*.*” (FG1, P7)*

Residents disagreed on the utility of didactic education in learning about sustainability, with some noting that their schedules were already overburdened with didactic curricula and others believing that some didactics would be helpful in establishing a knowledge baseline.

> *“…adding on more and more topics that are taking away from our core education and didactics may not end up being effective. And so I really do like the idea of having [a curriculum] being integrated into a quality improvement [project]*.*” (FG1, P5)*
>
> *“*… *because we’re coming into this with such different levels of knowledge and understanding, I personally would love to have an hour lecture about just the facts. [Then] we can kind of go into, you know, whatever sustainability work or improvements we want to make with a shared understanding…” (FG1, P6)*

Moreover, some residents suggested that various aspects of the curriculum should be tailored to resident levels. For example, a general review on climate change and human health may be most appropriate for an intern, while a focused review on the waste impact of a particular supply may be especially relevant to a chief resident who will soon be making their own preference cards.

> *“[*…*] having optional avenues for pursuing further education [could be] more targeted towards PGY level. [*…*] Best practices and more of the nuanced detail maybe not be as appropriate for a broader audience who may have varying views or backgrounds on the topic*.*” (FG3, P2)*

## Discussion

This study revealed that residents lack formal curricula on sustainability in surgery. While most residents agreed that sustainability was an important topic to learn, current knowledge was limited and variable. The few residents who were exposed to perioperative sustainability either received informal education on this topic, discovered it through an adjacent frame such as global surgery, or pursued independent research out of personal interest. Residents anticipated several barriers to implementing sustainable changes within the current context of academic surgery. Finally, the majority preferred practical education on implementing sustainable practices delivered through interactive formats such as quality improvement projects. On the other hand, some felt that some level-specific didactic content would be helpful to establish a knowledge base.

Many of our study’s thematic conclusions are in line with previous surgical curricular needs assessments, which frequently suggest that specialty curricula first require a basic foundational knowledge of a subject, and thereafter can be differentiated by trainee level.^11^ Prior needs assessment studies suggest that surgical culture and institutional structure often pose barriers to the implementation of new surgical practices, which need to be addressed within the curricula.^12^ While these conclusions are similar to the themes presented in this paper, it is important to note that this study’s thematic analysis is not directly comparable to others, given that there are no published needs assessments specifically addressing curricula on surgical sustainability. However, within related healthcare fields and at the pre-clinical level, it is well established that lack of knowledge and skill is the leading barrier to environmental stewardship in healthcare.^6,12^ As suggested in this paper, combating this lack of knowledge by integrating sustainability educational objectives into pre-clinical and clinical training is paramount.^13,14^

It is important to ask whether a surgical sustainability curriculum is necessary, as the absence of one does not necessarily justify its need. Furthermore, focus group participants noted that prior curriculum efforts were not always well received given the time constraints of surgical training. However, we believe that a sustainability curriculum is essential despite these constraints for the following reasons. First, the Accreditation Council for Graduate Medical Education (ACGME) lists “systems based practice” as a core competency for resident training. Climate change will affect patients, providers, and the healthcare infrastructure, all of which comprise the healthcare system.^3^ Second, we believe that a practical sustainability curriculum will push residents to develop into better surgeons. The majority of surgery-associated emissions come from the extensive single-use supply chain that pervades most of surgical care today. Interventions that improve the sustainability of surgical practice often involve switching from single-use supplies to reusable alternatives (or forgoing their use altogether). As such, by adopting sustainable practices, surgeons may become more efficient, learn how to do operations using fewer supplies, and reduce the cost of their practices.

This study has several limitations. First, our study is susceptible to selection bias as participants voluntarily signed up for the focus groups. Thus, participants with a baseline interest in sustainability may have been more likely to attend. As such, our results may overestimate both resident background knowledge and interest in a sustainability curriculum. Similarly, most participants were from UCSF, an academic medical center that already prioritizes surgical sustainability and is in a region with a culture of environmentalism.^15^ Thus, our results may be susceptible to institutional and geographic biases that similarly overestimate the perceived interest in sustainability. We attempted to mitigate this bias by hosting a virtual focus group that included participants from other parts of the U.S., including the Midwest, South, and Mid-Atlantic regions. Lastly, most focus group participants were general surgery residents, and there was less representation of surgical subspecialty residents. However, this should not significantly bias the results, as the principles of operating room waste are subspecialty agnostic, and our goal is that this curriculum will be relevant to all surgical specialties.

Based on our needs assessment, we envision a three-part curriculum. The first and foundational section of the curriculum would be a series of self-paced didactic modules about sustainability in surgery covering topics such as the science of climate change, how surgical care generates waste that leads to climate change, and a review of current initiatives to improve sustainability in surgery. These didactic modules would provide foundational knowledge. The second part of the curriculum would be a series of case studies highlighting actual sustainability improvement projects. These case studies would highlight project successes, implementation barriers, and failures. The third part of the curriculum would be a longitudinal quality improvement experience through which residents could guide or contribute to sustainability-focused quality improvement (QI) projects for their residency program. Completion of QI projects would subsequently generate new case studies. These three components would address the unmet need for a foundational sustainability curriculum while simultaneously allowing residents to tailor their experience to their interest level and availability. Future efforts will include developing parallel curricula for other perioperative stakeholders such as anesthesia residents, operating room nurses, and surgical technologists.

This study is the first of its kind addressing the need for a sustainability curriculum for surgical residents. It identifies a gap in baseline knowledge and highlights resident perspectives regarding how the curriculum should be structured. This study has both immediate and long-term implications. In the short term, a curriculum resulting from this work aims to grant trainees the knowledge to make informed decisions about their operating room resource utilization whenever possible, contextualize their choices within the broader hospital system, and understand how patients are impacted by climate change. In the long term, these efforts will empower surgeons to make sustainable choices in the operating room and educate future generations of surgeons. It is our hope that such efforts ultimately contribute to carbon footprint reduction.

## Data Availability

All data produced in the present study are available upon reasonable request to the authors.

## Notes

**Disclosure** The authors report no financial conflicts of interest related to this work.

### Competing Interest Statement

The authors have declared no competing interest.

### Funding Statement

This study did not receive any funding.

### Author Declarations

This study was approved by the University of California, San Francisco's (UCSF) Institutional Review Board (IRB 23-39152).

